# Increased angiotensin-converting enzyme 2, sRAGE and immune activation, but lowered calcium and magnesium in COVID-19: association with chest CT abnormalities and lowered peripheral oxygen saturation

**DOI:** 10.1101/2021.03.26.21254383

**Authors:** Hussein Kadhem Al-Hakeim, Hawraa Kadhem Al-Jassas, Gerwyn Morris, Michael Maes

**Affiliations:** Department of Chemistry, College of Science, University of Kufa, Iraq; Department of Pharmaceutical Chemistry, Faculty of Pharmacy, University of Kufa, Iraq; School of Medicine, IMPACT-the Institute for Mental and Physical Health and Clinical Translation, Deakin University, Barwon Health, Geelong, Australia; Department of Psychiatry, Medical University of Plovdiv, Plovdiv, Bulgaria; Department of Psychiatry, Faculty of Medicine, Chulalongkorn University, Bangkok, Thailand

**Keywords:** COVID-19, sRAGE, ACE2, inflammation, immune, oxidative stress, IL-6, IL-10, biomarkers

## Abstract

**Background:** The characterization of new biomarkers of COVID-19 is extremely important. Few studies measured the soluble receptor for advanced glycation end product (sRAGE), angiotensin-converting enzyme 2 (ACE2), calcium and magnesium in COVID-19.

**Aims:** To measure sRAGE, ACE2, interleukin (IL) -6, IL-10, CRP, calcium, magnesium, and albumin in COVID-19 patients in association with peripheral oxygen saturation (SpO2) and chest CT scan abnormalities (CCTA) including ground glass opacities.

**Methods:** This study measured sRAGE, ACE2, IL-6, IL-10, CRP using ELISA techniques, and calcium, magnesium, and albumin using a spectrophotometric method in 60 COVID-19 patients and 30 healthy controls.

**Results:** COVID-19 is characterized by significantly increased IL-6, CRP, IL-10, sRAGE, ACE2, and lowered levels of SpO2, albumin, magnesium and calcium. Neural networks showed that a combination of calcium, IL-6, CRP, and sRAGE yielded an accuracy of 100% in detecting COVID-19 patients with calcium being the most important predictor followed by IL-6, and CRP. COVID-19 patients with CCTAs showed lower SpO2 and albumin levels than those without CCTAs. SpO2 was significantly and inversely correlated with IL-6, IL-10, CRP, sRAGE, and ACE2, and positively with albumin, magnesium and calcium. Patients with positive IgG results showed a significant elevation in the serum level of IL-6, sRAGE, and ACE2 compared to the negatively IgG patient subgroup.

**Conclusion:** The results show that immune-inflammatory and RAGE pathway biomarkers may be used as external validating criterion for the diagnosis COVID-19. Those pathways coupled with lowered SpO2, calcium and magnesium are drug targets that may help to reduce the consequences of COVID-19.

## Introduction

Severe acute respiratory syndrome coronavirus-2 (SARS-CoV-2)-induced coronavirus disease-19 (COVID-19) is one of the biggest threats of our time (Azkur *et al*., 2020). The clinical spectrum of SARS-CoV-2 infection tends to be very wide, ranging from asymptomatic infection, minor illness to moderate upper respiratory tract disease to serious viral pneumonia with respiratory failure and death (Song *et al*., 2020, Zhou *et al*., 2020b). SARS-CoV-2 may invade the pulmonary cells and damage lung tissues (Xu *et al*., 2020). Although the quantitative rRT-PCR has been identified as the gold-standard for COVID-19 diagnosis, there are many difficulties, including the high cost and in its use because the test is time-consuming and needs availability of instruments and trained staff (Oliveira *et al*., 2020). Serological assays of SARS-CoV-2 antibodies are relatively easier to perform, but these assays show lower diagnostic performance, for example, because antibodies occur later in the disease (Zainol Rashid *et al*., 2020). Chest computed tomography and the assessment of ground-glass opacities, consolidation, crazy-paving patterns is of key importance for the early detection and prognosis and evaluation of the severity of COVID-19 pneumonia (Dai *et al*., 2020, Pan *et al*., 2020). Chest CT abnormalities (CCTA) have been reported in more than 70% of RT-PCR test–proven COVID-19 cases, including ground-glass opacities (GGOs), vascular enlargement, bilateral abnormalities, lower lobe involvement, and posterior predilection (Adams *et al*., 2020). Peripheral oxygen saturation (SpO2, with a normal range of 92%-98% (Shenoy *et al*., 2020), is often decreased in COVID-19 and especially in the more severe cases and stages of CCTAs (Dai *et al*., 2020, Luks and Swenson, 2020).

Due to high morbidity and mortality and lack of adequate treatments for seriously ill patients (Guan *et al*., 2020b, MacLaren *et al*., 2020), early identification and prediction of prognosis are crucial. Therefore, the development of biomarker tools to confirm the diagnosis and estimate the prognosis of SARS-CoV-2 infection, and to discover new drug targets to treat the consequences of the infection are an important field of study.

Recent studies showed increased levels of angiotensin-converting enzyme 2 (ACE2) following SARS-CoV-2 infection (Patel *et al*., 2021), and a case report showed very high ACE2 levels in a critically ill COVID-19 patient (Nagy *et al*., 2021). ACE2 is a common binding site for both SARS-CoV and SARS-CoV-2 (Zhu *et al*., 2018, Hoffmann *et al*., 2020). COVID-19 virus binds to ACE2 receptors in human cells through Spike proteins-S homotrimers with high affinity and leads mainly to endocytosis through the pathway of “ clathrin-mediated endocytosis (Amini Pouya *et al*., 2020). After attachment, transmembrane serine protease 2 (TMPRSS2) and several lysosomal proteases cleave and activate the S-glycoprotein due to SARS-CoV-2 entry into the cell through endocytosis or direct membrane fusion with the host membrane (Vlachakis *et al*., 2020). sACE2 is cleaved from ACE2 by a disintegrin and metallopeptidase domain-17 (ADAM17) and then released into the extracellular environment (Lambert *et al*., 2005).

SARS-CoV-2 may cause an overzealous immune-inflammatory response and even a cytokine storm in the host associated with severe lung pathology (Hui and Zumla, 2019, Huang *et al*., 2020a). These processes are closely linked to poor clinical outcomes (Huang *et al*., 2020b, Qin *et al*., 2020), worsening of hypercoagulation (Mehta *et al*., 2020), and higher mortality (Wu *et al*., 2020). For example, the severity of COVID-19 infection is associated with increased interleukin-6 (IL-6) and other proinflammatory cytokines, C-reactive protein (CRP), and IL-10, a negative immune-regulatory cytokine (Ruan *et al*., 2020, Wu *et al*., 2020). Mitigation of the cytokine storm may contribute to better clinical outcomes in severe cases of COVID-19. (Ye *et al*., 2020). Severe COVID-19 is also accompanied by lowered serum albumin levels or hypoalbuminemia as part of the systemic inflammatory response in patients with COVID-19 (Huang *et al*., 2020c) (Zhang *et al*., 2020c, Zhou *et al*., 2020b). Hypoalbuminemia frequently occurs in inflammatory diseases because the liver production of albumin is downregulated by pro-inflammatory cytokines, including IL-6 (Maes, 1993), and because capillary permeability is enhanced leads to leakage of albumin to the interstitial space (Soeters *et al*., 2019).

Another possible candidate biomarker of COVID-19 is a soluble form of the advanced glycation end-product receptor (sRAGE) (Yalcin Kehribar *et al*., 2021). Advanced glycation end products (AGEs) and high mobility group box 1 protein (HMGB1) can bind to the membrane-bound RAGE (mRAGE), thereby initiating intracellular signaling (Vistoli *et al*., 2013), which leads to activation of pro-inflammatory transcription factors, including nuclear factor (NF)-kappa B (Macaione *et al*., 2007, Julio *et al*., 2014), which in turn increased the production of, for example, IL-6 protein transcription in immune cells (Wang and Liu, 2016). sRAGE is generated through proteolytic cleavage of the RAGE extracellular domain or through alternative RNA splicing (Zhang *et al*., 2008b, Sterenczak *et al*., 2009).

Several studies have documented a high prevalence of hypocalcemia in COVID-19 patients (Cappellini *et al*., 2020, Lippi *et al*., 2020), and one study found that hypocalcemia could predict hospitalization in COVID-19 patients (Di Filippo *et al*., 2020). Alterations of Ca2+ homeostasis may occur during viral infections (Nieto-Torres *et al*., 2015) (Deng *et al*., 2012) because viruses often utilize Ca^2+^ signals to create a suitable cellular environment that meets their demands (Zhou *et al*., 2009). Lowered levels of calcium may interfere with calcium-regulated processes, including signal transduction networks, which are regulated by calcium’s activities as a second messenger or as plasma membrane channels and pumps (Civitelli and Ziambaras, 2011, Görlach *et al*., 2015). Magnesium deficiency is significantly and inversely related to the severity of infection in COVID-19 patients (Quilliot *et al*., 2020). Magnesium shows anti-inflammatory (Abiri and Vafa, 2020) and antioxidant (Güzel *et al*., 2019) properties and is an important cofactor for ATP, which mediates several basic enzymatic reactions (Romani and Scarpa, 1992). Nevertheless, no studies in SARS-CoV-2-infected patients with COVID-19 have examined whether a combination of the above biomarkers may be used to externally validate the diagnosis of COVID-19 and whether these pathways are associated with the presence of lowered SpO2.

Hence, the present study aimed to examine the levels of ACE2, sRAGE, IL-6, IL-10, albumin, calcium, and magnesium in patients with COVID-19 versus normal controls and to examine the association between those biomarkers and SpO2 levels and CCTAS. The specific hypotheses are that a) COVID-19 is associated with increased ACE2, sRAGE, IL-6, IL-10 levels, and lowered albumin, calcium, and magnesium ; and b) that those biomarkers are associated with lowered SpO2.

## Subjects and Methods

### Subjects

Sixty COVID-19 male patients aged 25-59 years were recruited at the Al-Sadr Teaching Hospital and Al-Amal Specialized Hospital for Communicable Diseases in Najaf governorate-Iraq between September and November 2020. All patients were admitted to these two centers, which are official quarantine centers specialized in the treatment of COVID-19. The diagnosis was made by senior physicians and virologists. All patients suffered from acute respiratory syndrome and were diagnosed with SARS-CoV-2 infection according to positive results of COVID-19 nucleic acids by reverse transcription real-time polymerase chain reaction (rRT-PCR), positive IgM, in addition to the standard symptoms of the disease including fever, breathing difficulties, cough, and loss of smell and taste. We excluded patients with premorbid medical illnesses, including diabetes type 1, liver, kidney and cardiovascular diseases. We also recruited 30 healthy controls, age, and sex-matched to the patient groups. All controls were free from any systemic disease. However, to enhance their immunity against COVID-19 infection, some healthy controls were taken zinc and vitamins C and D.

The “institutional ethics board of the University of Kufa” approved the study (617/2020). All participants gave written informed consent before participation in this study. The study was conducted according to Iraq and international ethics and privacy laws and was conducted ethically in accordance with the World Medical Association Declaration of Helsinki. Furthermore, our IRB follows the International Guideline for Human Research protection as required by the Declaration of Helsinki, The Belmont Report, CIOMS Guideline and International Conference on Harmonization in Good Clinical Practice (ICH-GCP).

#### Measurements

The RT-PCR tests were performed using the Applied Biosystems^®^ QuantStudio™ 5 Real-Time PCR System (Thermo Fisher Scientific) supplied by Life Technologies Holdings Pte Ltd., Marsiling Industrial Estate, Singapore. The Lyra^®^ Direct SARS-CoV-2 Assay kits were supplied by Quidel Corporation, CA, USA. The procedures were exactly followed as mentioned by the manual of the kits. This is a real-time RT-PCR assay for the qualitative detection of human coronavirus SARS-CoV-2 from viral RNA extracted from nasal, nasopharyngeal or oropharyngeal swab specimens. The Assay targets the non-structural Polyprotein (pp1ab) of the SARS-CoV-2 virus. Patients had a chest X-ray and chest computed tomography scan (CT-scan) to search for lung abnormalities. CT-scans were made by using SOMATOM Definition AS, Supplied by Seimens, Munchen, Germany. The chest CT abnormalities (CCTAs) assessed in our study comprise GGOs, areas of pulmonary densification consistent with residual lesions, pneumonic consolidation, and crazy-paving patterns (Kwee and Kwee, 2020). The total CCTA score was computed using the international standard nomenclature (Hansell et al., 2008; Franquet, 2011). Accordingly, the patients’ group was subdivided into two subgroups depending on the presence of CCTAs into those with (COVID9+CCTA) and without (COVID-CCTA).

The blood samples were taken in the early morning between 7.30-9.00 a.m. after awakening and before having breakfast. Five milliliters of venous blood samples were drawn and transferred into clean plain tubes. Hemolyzed samples were rejected. After ten minutes, the clotted blood samples were centrifuged for five minutes at 3000 rpm and then serum was separated and transported into three new Eppendorf tubes until assay. A qualitative ACON^®^ COVID-19 IgG/IgM rapid test was used to detect IgG and IgM in the sera of patients and controls. The kits have a sensitivity ≥ 99.1 % and a specificity ≥ 98.2 %. We also divided the patient’s subgroups according to IgG results into negative-IgG and positive-IgG subgroups to examine the difference in the measured biomarkers between these subgroups. A C-Reactive Protein (CRP) latex slide test (Spinreact^®^, Barcelona, Spain) was used for the qualitative and semi-quantitative CRP measurement in human serum. The serum titer is the reciprocal of the maximum dilution exhibiting a positive response multiplied by a positive control concentration. The approximate CRP concentration in the patient sample is calculated as follows: 6 x CRP Titer = mg/L. Serum IL-6, IL-10, sRAGE, and sACE2 were measured using ELISA kits supplied by Melsin Medical Co, Jilin, China. All kits were based on a sandwich technique and showed an inter-assay CV of less than 12%. Total calcium, albumin, and magnesium were measured spectrophotometrically by kits supplied by Biolabo^®^, Maizy, France. The procedures were followed according to the manufacturer’s instructions and without modifications.

### Statistical analysis

Analysis of variance (ANOVA) was used to check differences in scale variables among groups, and analysis of contingency tables (χ^2^-test) was used to assess associations between nominal variables. We computed correlation matrices based on Pearson’s product-moment correlation coefficients to determine correlations between biomarkers and clinical scores (e.g., duration of illness and SpO2). We used univariate generalized linear model (GLM) analysis to delineate the associations between diagnosis and the biomarkers while controlling for confounding variables, including tobacco use disorder (TUD), age, and BMI. The effect size was estimated using partial eta-squared values. We also computed model generated (GLM analysis) estimated marginal mean (SE) values and protected pairwise comparisons among treatment means. Multiple comparisons were evaluated using false discovery rate p-correction (Benjamini and Hochberg, 1995). Tests were 2-tailed, and a p-value of 0.05 was used for statistical significance.

Multilayer perceptron Neural Network models were employed to assess the more complex associations between the diagnosis of COVID-19 *versus* controls (entered as output variables) and biomarkers (entered as input variables). We trained the models with an automated feed-forward architecture, two hidden layers with up to 4 nodes in each layer, with minibatch training with gradient descent, and 20-50 epochs. The stopping rule was one consecutive step with no further decrease in the error term. Three samples were extracted, namely “a training sample to estimate the network parameters (46.67% of all participants), testing set to prevent overtraining (20.0%) and a holdout set to evaluate the final network (33.33%). Error, relative error, and importance and relative importance of all input variables were computed” (Moustafa *et al*., 2020). All statistical analyses were performed using IBM SPSS windows version 25, 2017.

## Results

### Socio-demographic data

**Table 1** demonstrates the socio-demographic data in COVID+CCTA and COVID-CTTA patients, and healthy controls (HC). No significant differences among these study groups were detected in BMI, education, residency, marital status, employment, and TUD. CCTA+ patients were somewhat older than CCTA-patients, but illness duration was not significantly different between those groups. All patients were on O2 therapy and were treated with paracetamol, bromhexine, vitamin C, vitamin D, and zinc as a continuous treatment. There were no significant differences in the use of azithromycin, enoxaparin, dexamethasone, famotidine, heparin, and meropenem between CCTA+ and CCTA-patients.

**Table 1.** Socio-demographic and clinical data of COVID-19 patients divided into those with (COVID+CCTA) and without (COVID-CCTA) chest CT scan abnormalities, and healthy controls (HC).

| Variables | HC<br>(n=30) | COVID-CCTA<br>(n=13) | COVID+CCTA<br>(n=47) | F/FEPT/$\chi^2$ | df | p |
| --- | --- | --- | --- | --- | --- | --- |
| Age (years) | 47.1±5.2 | 42.92±10.1<sup>B</sup> | 50.1±7.4<sup>C</sup> | 5.47 | 2/87 | 0.006 |
| BMI (kg/m<sup>2</sup>) | 28.1±2.2 | 29.8±5.8 | 27.8±4.5 | 1.32 | 2/87 | 0.273 |
| Education, years | 11.3±4.5 | 12.5±3.43 | 10.9±3.6 | 0.84 | 2/87 | 0.436 |
| Duration of illness (months) | 14.5±6.5 | 14.3±4.7 | 14.4±5.0 | 0.02 | 1/58 | 0.891 |
| Urban/Rural | 27/3 | 10/3 | 33/14 | 4.16 | 2 | 0.125 |
| Single/married | 28/2 | 11/2 | 43/4 | 0.87 | 2 | 0.648 |
| TUD (No/Yes) | 10/20 | 5/8 | 16/31 | 0.11 | 2 | 0.945 |
| Employment (No/Yes) | 8/22 | 8/5 | 9/38 | FEPT | - | <0.001 |
| Zinc (No/Yes) | 16/14 | 0/13 | 0/47 | FEPT | - | <0.001 |
| Vitamin D (No/Yes) | 25/5 | 0/13 | 0/47 | FEPT | - | <0.001 |
| Vitamin C (No/Yes) | 16/14 | 0/13 | 0/47 | FEPT | - | <0.001 |
| Dexamethasone (No/Yes) | - | 3/10 | 6/41 | 0.85 | 1 | 0.357 |
| Famotidine (No/Yes) | - | 1/12 | 22/25 | FEPT | - | 0.011 |
| Azithromycin (No/Yes) | - | 5/8 | 20/27 | 0.07 | 1 | 0.791 |
| Meropenem (No/Yes) | - | 8/5 | 27/20 | 0.07 | 1 | 0.791 |
| Heparin (No/Yes) | - | 5/8 | 20/27 | 0.07 | 1 | 0.791 |
| Enoxaparin (No/Yes) | - | 8/5 | 27/20 | 0.07 | 1 | 0.791 |
| O2 therapy (No/Yes) | - | 0/13 | 0/47 | - | - | - |
| Bromhexine (No/Yes) | - | 0/13 | 0/47 | - | - | - |
| Paracetamol (N/Yes) | - | 0/13 | 0/47 | - | - | - |

All results are shown as mean (SD). FEPT: Fisher's exact probability test. BMI: body mass index, TUD: tobacco use disorder.

### Differences in biomarkers between CCTA+ and CCTA-patients

**Table 2** shows the different biomarkers’ measurements in the three study groups with significant differences in all biomarkers, which remained significant after p-correction for FDR. There was a significant decrease in S pO2 and serum albumin in CCTA+ patients compared to CCTA-patients. The serum levels of IL-6, IL-10, sRAGE, sACE2, and CRP were higher in COVID-19 patients compared with controls. SpO2, serum albumin, calcium, and magnesium were lowered in both patient groups compared to controls, while there were no significant differences in those biomarkers between both patients’ subgroups.

**Table 2.** Biomarkers in COVID-19 patients divided into those with (COVID+CCTA) and without (COVID-CCTA) chest CT scan abnormalities, and healthy controls (HC).

| **Biomarkers** | **HC**<br>**(n=30)** | **COVID-CCTA**<br>**(n=17)** | **COVID+CCTA**<br>**(n=43)** | **F** | **p** | **Partial**<br>**$\eta^2$** |
| --- | --- | --- | --- | --- | --- | --- |
| SPO<sub>2</sub> | 97.1 ± 1.4<sup>B, C</sup> | 81.9 ± 12.5<sup>A, C</sup> | 69.8 ± 12.2<sup>A, B</sup> | 64.97 | <0.001 | 0.607 |
| IL-6 pg/ml | 9.5 ± 3.2<sup>B, C</sup> | 14.7 ± 2.6<sup>A</sup> | 14.7 ± 3.5<sup>A</sup> | 26.94 | <0.001 | 0.391 |
| CRP mg/dL | 2.5 ± 0.0<sup>B, C</sup> | 5.3 ± 3.4<sup>A</sup> | 7.9 ± 5.3<sup>A</sup> | 25.45 | <0.001 | 0.377 |
| Albumin g/l | 44.8 ± 5.0<sup>B, C</sup> | 38.7 ± 6.8<sup>A, C</sup> | 33.5 ± 5.6<sup>A, B</sup> | 34.60 | <0.001 | 0.452 |
| IL-10 pg/ml | 5.5 ± 4.3<sup>B, C</sup> | 14.6 ± 5.5<sup>A</sup> | 14.6 ± 4.9<sup>A</sup> | 55.90 | <0.001 | 0.571 |
| sRAGE pg/ml | 456.2 ± 232.0<sup>B, C</sup> | 967.6 ± 202.0<sup>A</sup> | 928.3 ± 202.6<sup>A</sup> | 69.23 | <0.001 | 0.622 |
| sACE2 ng/ml | 2.71 ± 1.34<sup>B, C</sup> | 4.19 ± 1.24<sup>A</sup> | 4.05 ± 1.19<sup>A</sup> | 11.71 | <0.001 | 0.218 |
| Magnesium mM | 0.76 ± 0.15<sup>B, C</sup> | 0.64 ± 0.08<sup>A</sup> | 0.64 ± 0.15<sup>A</sup> | 6.92 | 0.002 | 0.142 |
| Calcium mM | 2.50 ± 0.22<sup>B, C</sup> | 2.00 ± 0.16<sup>A</sup> | 1.89 ± 0.28<sup>A</sup> | 55.93 | <0.001 | 0.571 |

All results are shown as mean (SE). All results of univariate GLM analyses examining the associations between diagnosis (healthy control and COVID-19 patients divided into two groups and after adjusting for age, tobacco use disorder, and body mass index; all df=2/84.
IL: interleukin, CRP: C-reactive protein, sRAGE: soluble receptor for advanced glycation end products, sACE2: soluble Angiotensin-converting enzyme 2, SPO<sub>2</sub> %: peripheral oxygen saturation percentage.

### Neural network results

We have performed neural network analyses with diagnosis (COVID-19 *versus* controls) as output variables and the biomarkers as explanatory variables and reran the analyses with the 4 most important biomarkers, i.e., calcium, IL-6, CRP, and sRAGE. **Table 3** shows the network information of the model examining the separation of COVID-19 versus controls. The network has been trained using two hidden layers with three and two units in layers 1 and 2. Hyperbolic tangent was used as the activation function in hidden layer 1 and identity in the output layer. **Table 3** displays the partitioned confusion matrices showing an AUC ROC=1.000 with an accuracy of 100.0% in the holdout sample with a sensitivity of 100.0% and a specificity of 100.0%. **Figure 1** shows the relative importance of the most important input variables (Ca > IL-6 > CRP > sRAGE), representing the most important determinants of the predictive power of the model.

**Table 3.**
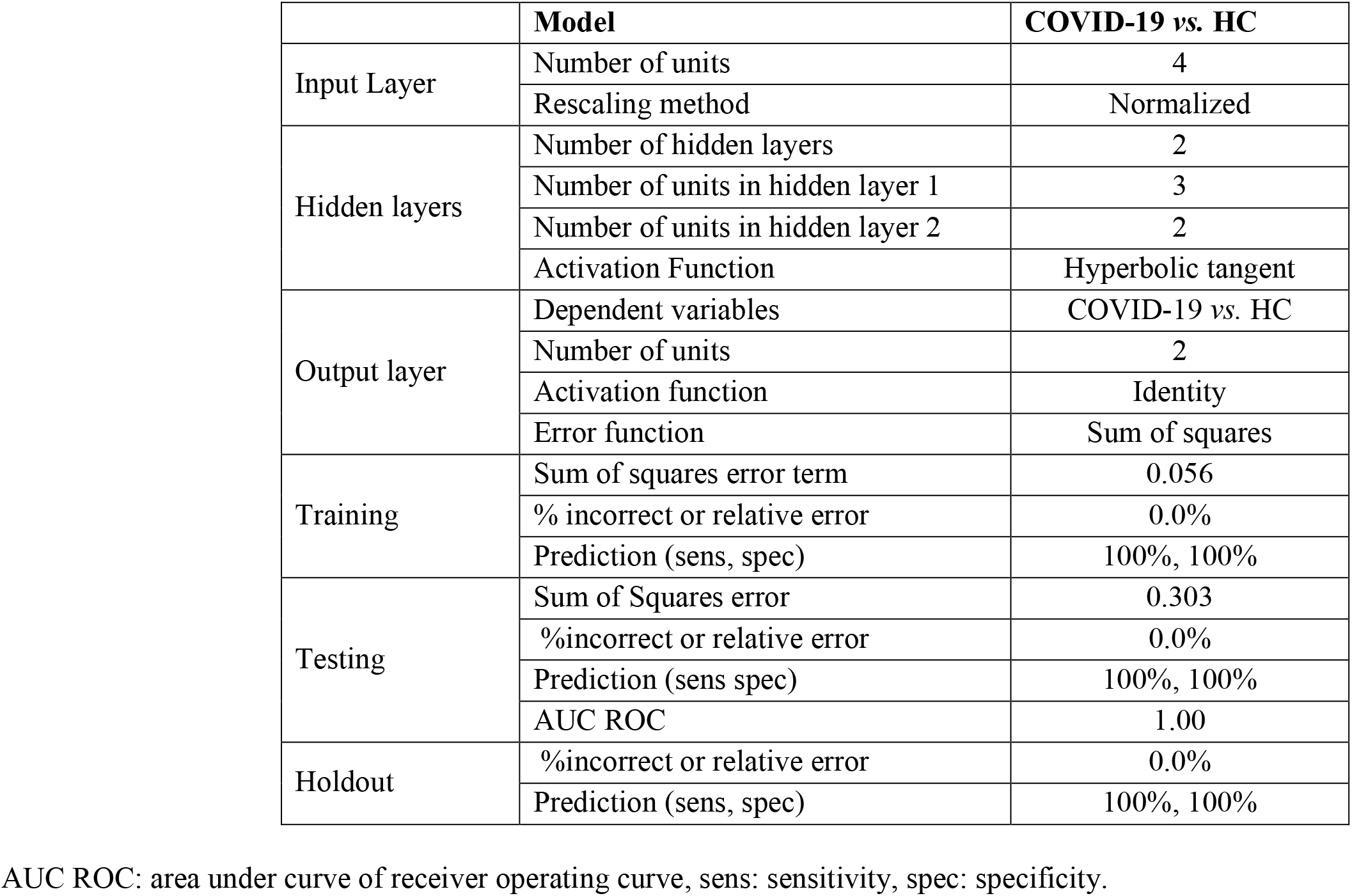
Results of neural networks with diagnosis with COVID-19 patients versus heathy controls (HC) as output variables and biomarkers as input variables.

**Table 4.** Differences in biomarkers between COVID-19 patients with and without anti-SARS-CoV-2 IgG antibodies

| Parameter | Negative IgG<br>N=20 | Positive IgG<br>N=40 | F | df | p | Partial $\eta^2$ |
| --- | --- | --- | --- | --- | --- | --- |
| SPO<sub>2</sub> % | 69.8 ± 2.9 | 73.8 ± 2.1 | 1.26 | 1 | 0.266 | 0.021 |
| IL-6 pg/ml | 13.3 ± 0.7 | 15.4 ± 0.5 | 6.47 | 1 | 0.014 | 0.100 |
| CRP mg/dL | 6.8 ± 1.1 | 7.6 ± 0.8 | 0.01 | 1 | 0.909 | <0.001 |
| Albumin g/l | 34.2 ± 1.4 | 34.9 ± 1.0 | 0.15 | 1 | 0.705 | 0.002 |
| IL-10 pg/ml | 13.6 ± 1.1 | 15.0 ± 0.8 | 1.72 | 1 | 0.194 | 0.029 |
| sRAGE pg/ml | 833.2 ± 42.3 | 988.6 ± 29.9 | 10.55 | 1 | 0.002 | 0.154 |
| sACE2 ng/ml | 3.28 ± 0.24 | 4.48 ± 0.17 | 17.12 | 1 | <0.001 | 0.228 |
| Magnesium mM | 0.66 ± 0.03 | 0.62 ± 0.02 | 1.08 | 1 | 0.303 | 0.018 |
| Calcium mM | 1.87 ± 0.05 | 1.93 ± 0.04 | 0.59 | 1 | 0.446 | 0.010 |

All results of univariate GLM analysis; results are shown as mean ± SE. IL: interleukin, CRP: C-reactive protein, sRAGE: soluble receptor for advanced glycation end products, sACE2: soluble angiotensin-converting enzyme 2, SPO<sub>2</sub> %: peripheral oxygen saturation percentage.

**Figure 1.**
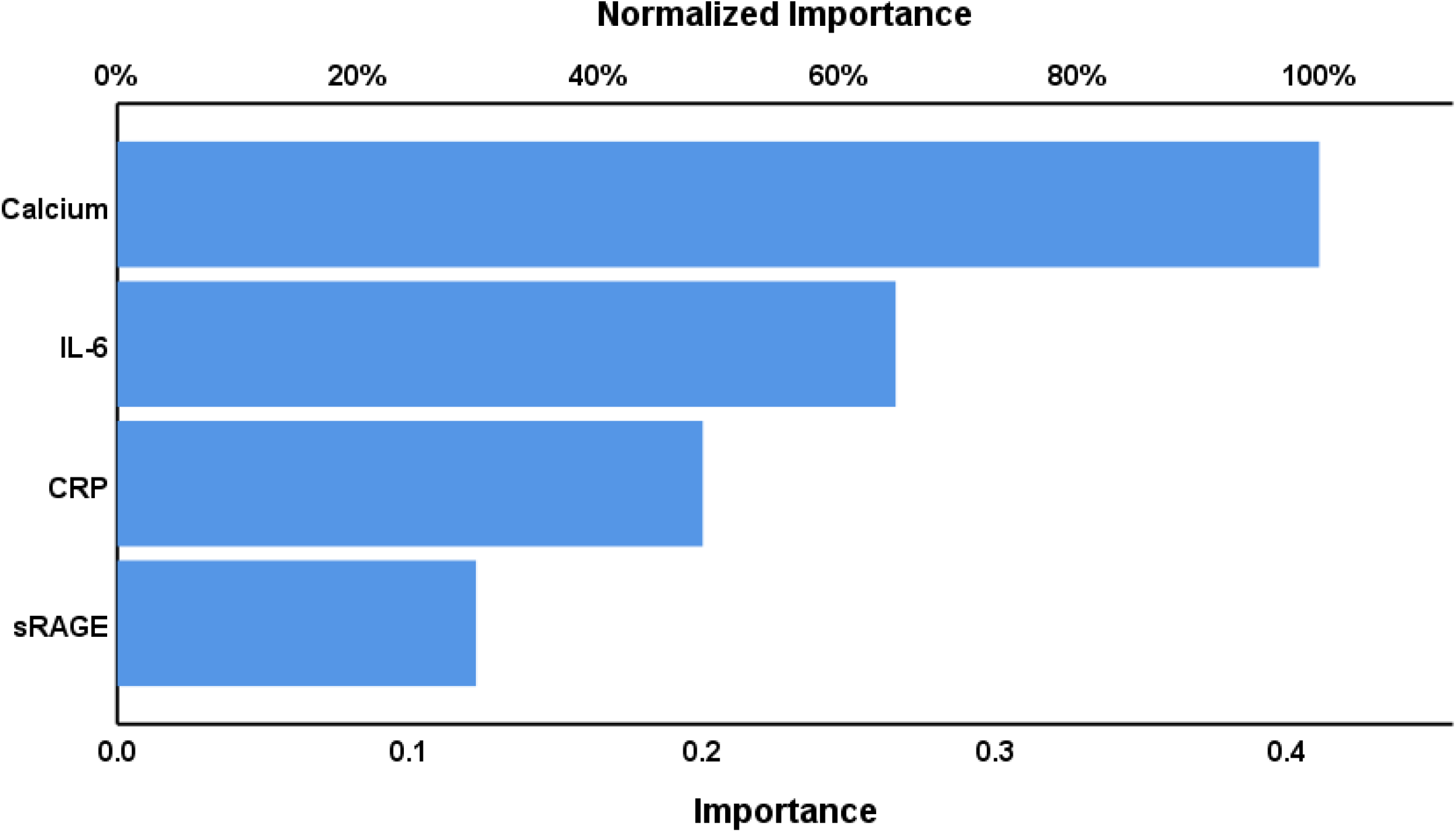
Results of neural network (importance chart) with diagnosis of COVID-19 as output variables and biomarkers (in z-scores) as input variables. IL-6: interleukin-6, CRP: C-reactive protein, sRAGE: soluble receptor for advanced glycation end product.

### Biomarkers results in COVID-19 subgroups with and without anti-SARS-CoV-2 IgG antibodies

The univariate GLM analysis of the biomarkers in patients’ subgroups (positive versus negative anti-SARS-CoV-2 IgG antibodies showed significant elevations in serum IL-6, sRAGE, and ACE2 in patients with positive IgG results as compared with the IgG negative patient subgroup. These differences remained significant after performing p-correction for FDR, even IL-6 (p=0.042). Other biomarkers showed no significant differences between those subgroups.

### Intercorrelation matrix

**Table 5** shows the intercorrelation matrix of the biomarkers and duration of the infection and SpO2 levels. Duration of illness was inversely associated with SpO2, albumin, magnesium, and calcium. These p-values remained significant after FDR correction (all at p<0.003). The results showed that SpO2 was significantly and negatively correlated with duration of illness, CRP, IL-6, IL-10, sRAGE, and ACE2, and significantly and positively with albumin, magnesium, and calcium. There were no significant associations between the biomarkers and the total CCTA score.

**Table 5.** Intercorrelation matrix between biomarkers and duration of COVID-19 infection and peripheral oxygen saturation (SpO_2_)

| Parameter | Duration of infection | SpO<sub>2</sub> |
| --- | --- | --- |
| Duration of Disease | - | -0.410 (0.001) |
| SPO<sub>2</sub> | -0.410 (0.001) | - |
| IL-6 | 0.007 (0.958) | -0.444 (<0.001) |
| CRP | 0.152 (0.248) | -0.638 (<0.001) |
| IL-10 | -0.051 (0.699) | -0.492 (<0.001) |
| sRAGE | 0.097 (0.461) | -0.461 (<0.001) |
| sACE2 | 0.106 (0.419) | -0.247 (0.019) |
| Albumin | -0.515 (<0.001) | 0.696 (<0.001) |
| Magnesium | -0.333 (0.009) | 0.469 (<0.001) |
| Calcium | -0.405 (0.001) | 0.745 (<0.001) |

Shown are Pearson's product moment correlation coefficients (p-value). IL: interleukin, CRP: C-reactive protein, sRAGE: soluble receptor for advanced glycation end products, sACE2: soluble angiotensin-converting enzyme 2, SPO<sub>2</sub> %: peripheral oxygen saturation percentage.

## Discussion

The first major finding of this study is that COVID-19 is accompanied by increased IL-6, IL-10, and CRP in addition to sRAGE and ACE2 and that albumin, calcium and magnesium are significantly decreased as compared with controls. Moreover, a combination of calcium, IL-6, CRP and sRAGE was able to validate the diagnosis of COVID-19 with 100% accuracy. Duration of infection was accompanied by lowered SpO2, albumin, calcium and magnesium levels.

### Inflammation and COVID-19

The pathophysiology of COVID-19 involves a potent immune-inflammatory response, including increases in IL-6 and IL-10, which is associated with a higher risk of disease deterioration (Han *et al*., 2020). When the immune system is activated following infection with SARS-CoV-2, an overzealous immune system may secrete many proinflammatory cytokines, and may cause a cytokine storm (Kuba *et al*., 2010, Jia, 2016), which is a key component in the pathophysiology of severe COVID-19 (Tetro, 2020, Yang *et al*., 2020) (Huang *et al*., 2020a, Li *et al*., 2020a). Sustained elevations in pro-inflammatory cytokines in COVID-19 patients indicate continuing systemic and tissue inflammation (Chen *et al*., 2020, Guan *et al*., 2020a, Wang *et al*., 2020). It is interesting to note that the combination of IL-6 coupled with D-dimer may be used to differentiate extreme COVID-19 cases (Gao *et al*., 2020).

Increased CRP levels have been associated with the cytokine storm and liver damage in COVID-19 patients and generally contribute to a worse prognosis (Li *et al*., 2020b). The hypoalbuminemia in our COVID-19 patients may, at least in part, be explained by the acute-phase response downregulating negative acute-phase proteins, although increased vascular permeability and liver or kidney disease may play a role (Ronit *et al*., 2020). Albumin has strong antioxidant properties (Maes *et al*., 2011), and as such, hypoalbuminemia may play a role in the exaggerated immune responses in COVID-19. Moreover, albumin has anticoagulant and antiplatelet activities, and therefore hypoalbuminemia may be associated with hypercoagulability (Paar *et al*., 2017), increased D-dimers, a thrombin generation marker (Thachil *et al*., 2020), and enhanced risk of artery and venous thrombosis (Ronit *et al*., 2020). It is important to note that coagulopathies complicate the clinical course of COVID-19 (Thachil *et al*., 2020) (Huang *et al*., 2020c) and that, therefore, hypoalbuminemia may play a role in COVID-19 associated coagulopathies.

### ACE2 and COVID-19

As described in the Introduction, only a few studies reported increased ACE2 levels in patients with SARS-CoV-2 infection and COVID-19 (Nagy *et al*., 2021, Patel *et al*., 2021). ACE2 concentrations are strongly related to ACE2 catalytic activity (Zhang *et al*., 2018). Increased ACE2 plasma levels are associated with an increased risk of stroke, myocardial infarction, incident diabetes, incident heart failure and, as a consequence, with total cardiovascular and non-cardiovascular deaths (Narula *et al*., 2020). In patients with heart failure, increased plasma ACE2 activity is associated with a worsened prognosis (Oudit and Pfeffer, 2020). The most important determinants of increased ACE2 values are male sex, geographic ancestry, and body-mass index (BMI) (Narula *et al*., 2020). In physiological conditions, cell-bound membrane ACE2 is a counter-regulatory pathway through the production of angiotensin-(1-7) and stimulation of the angiotensin 2 (AT2R) and Mas receptors, which promote anti-inflammatory and antioxidant processes, vasorelaxation, anti-fibrosis, and attenuate the renin-angiotensin system, cellular proliferation, hypertrophy, and vasoconstriction (Narula *et al*., 2020). Nevertheless, the extracellular domain of the transmembrane ACE2 facilitates entry for SARS-CoV-2 into the cells via interactions with the SARS-CoV spike protein (Walls *et al*., 2020, Zhou *et al*., 2020a). This process is followed by downregulation of surface ACE2 expression that may culminate in loss of angiotensin II effects leading to increased multisystem diseases, including lung, cardiovascular and renal disease (Oudit and Pfeffer, 2020). Moreover, the downregulation of ACE2 expression through the SARS-CoV spike protein causes shedding of soluble ACE, which is therefore a consequence of SARS-CoV infection (Glowacka *et al*., 2010). It is hypothesized that increased sACE2 shedding may neutralize the SARS-CoV2 virus at a distance from virus entry into the cells (Brest *et al*., 2021). All in all, COVID-19 infection may cause part of its detrimental effect via renin-angiotensin system overactivation and increased levels of sACE2.

### sRAGE and COVID-19

As described in the Introduction, increased levels of plasma sRAGE were reported previously (Yalcin Kehribar *et al*., 2021). In another study, sRAGE was significantly increased in COVID-19 patients with diabetes mellitus versus normal controls (Dozio *et al*., 2020a). Nevertheless, in our study, patients with diabetes mellitus were excluded indicating that the increased sRAGE concentrations in COVID-19 are not due to diabetes mellitus. These findings extend those of Dozio et al. (2020) who reported that in COVID-19 patients, the RAGE pathway may be modulated regardless of glycemic control (Dozio *et al*., 2020b). It is important to note that increased sRAGE in bronchoalveolar lavage and plasma indicates acute pulmonary injury and type 1 alveolar epithelial cell injury (Uchida *et al*., 2006). Nevertheless, in our study, sRAGE levels were not significantly increased in COVID-19 patients with CCTA than those without CCTA. sRAGE may function as a decoy receptor, which prevents binding of ligands to mRAGE, thereby negatively regulating immune-inflammatory responses (Yang *et al*., 2014, Oczypok *et al*., 2017), as observed during acute lung injury (Zhang *et al*., 2008a), the acute respiratory distress syndrome (ARDS) (Izushi *et al*., 2016), neutrophilic asthma, and COPD (Sukkar *et al*., 2012). In some diseases, administration of sRAGE may attenuate immune-inflammatory responses (Ekong *et al*., 2006, van Zoelen *et al*., 2011, Zhang *et al*., 2017).

### Calcium, magnesium and COVID-19

In our study, the most important biomarker of COVID-19 was lowered calcium. Low calcium is common in severe COVID-19 patients and is associated with the severity of illness (Sun *et al*., 2020), and lowered calcium levels may detect the most critical COVID-19 patients with high specificity (Yang *et al*., 2021). However, hypocalcemia is also detectable in COVID-19 patients with non-severe illness (Pal *et al*., 2020). In addition, patients with very low calcium values have an increased prevalence of acute respiratory distress syndrome (ARDS) (Sun *et al*., 2020). Interestingly, inflammation appears to be higher in COVID-19 patients with extremely low blood calcium levels compared to patients with normal calcium levels (Di Filippo *et al*., 2020, di Filippo *et al*., 2021). This is not an invariant finding, however, as hypocalcaemia is sometimes detectable in COVID-19 patients with non-severe illness rather arguing against inflammation as a universal cause of the phenomenon (Pal *et al*., 2020). Hypocalcemia causes an increased secretion of PTH, hypoproteinemia, hypomagnesemia (Kelly and Levine, 2013) and more detrimental effects as described in the Introduction.

SARS-CoV and MERS-CoV are known to disrupt intracellular calcium homeostasis to foster membrane fusion and increase infectivity (Millet and Whittaker, 2 018, Straus *et al*., 2019) and the hypocalcemia seen in patients with severe COVID-19 is also evident in patients following SARS-CoV infection (Booth, 2003). This suggests that there may be a common mechanism driving the development of calcium dyshomeostasis following beta coronavirus infections. Several mechanisms are involved in calcium dyshomeostasis mediated by a plethora of viruses following cell invasion including activation of Ca ^2+^ ATPase pumps, the transient receptor potential canonical (TRPC) channels with the receptor-operated Ca^2+^ currents (ROCs), and the store-operated calcium entry (SOCE) channels (Chen *et al*., 2019). The latter may be of particular interest as one potential mechanism explaining beta coronavirus-mediated calcium dyshomeostasis is that SOCE may be activated by endoplasmatic-reticulum (ER) stress (Chen *et al*., 2015, Zhang *et al*., 2020b), which is an invariant consequence of the replication strategies employed by these viruses (Fung and Liu, 2014, Morris *et al*., 2020a). The plausibility of SOCE activation as a vehicle to explain SARS-CoV2 mediated decreases in plasma calcium levels is increased by the presence of in vivo data confirming that SARS-CoV ORF8 and ORF3 have the capacity to activate SOCE (Hyser and Estes, 2015). Importantly, SOCE is activated in conditions of inflammation and oxidative stress (Nunes and Demaurex, 2014), which is characteristic of SARS-CoV-2 infection (Morris *et al*., 2020b). SOCE activation also upregulates nuclear factor-KB potentially resulting in a positive feedback mechanism amplifying inflammation, oxidative stress and calcium dyshomeostasis (Berry *et al*., 2018, Dresselhaus and Meffert, 2019).

Lowered serum magnesium is associated with increased thrombosis risk, decreased fibrinolysis, endothelial dysfunction, mitochondrial dysfunctions, increased inflammatory and oxidative stress, and increased fatty acid production (Çiçek *et al*., 2016, Zheltova *et al*., 2016, Gromova *et al*., 2018, Nielsen, 2018). Furthermore, in vivo studies have demonstrated that magnesium has antithrombotic properties and decreases mortality in induced pulmonary thromboembolism (Sheu *et al*., 2003).

### CCTA and consequences

The second major finding of the present study is that CCTAs are accompanied by lowered albumin and SpO2 levels and that the latter is associated with all biomarkers described above. Chest imaging, especially by computed tomography scan (CT-scan), is of great importance for the diagnosis, management, and follow-up of patients with COVID-19 infection (Fang, 2020, Zhang *et al*., 2020a). For example, the appearance of GGOs in COVID-19 indicates severe or persistent inflammation in the lungs, and more severe symptoms including bronchiolitis and pneumonia in both lungs, and lung fibrosis (Sadhukhan *et al*., 2020). These lung infection sites may cause the recruitment of various immune cells, and consequently, a pro-inflammatory response is mounted, including increased levels of IL-6 (Sadhukhan *et al*., 2020). Our results also indicate that lung lesions as detected with chest-CT scan may lead to decreased oxygenation. In fact, many of the patients included here (88.3%) showed hypoxemia as indicated by SpO2 < 92%. Moreover, hypoxia may induce inflammation (Eltzschig *et al*., 2014) and may upregulate ACE2 gene expression and protein levels in lung and kidney tissues, which may further contribute to the severity of COVID-19 (Shenoy *et al*., 2020). There are also reports that patients with higher GGO CT-scores have lower calcium levels and that SPO2 is correlated with serum calcium with those COVID-19 patients staying longer in ICU (Yang *et al*., 2021).

### Biomarkers and IgG positivity

Another important result of our study is that patients with positive IgG results showed higher IL-6, sRAGE and ACE2 levels as compared to those with negative IgG results. In our study, 66.7% of the COVID-19 patients showed IgG positivity which is in agreement with previous studies showing that around 77.9% of plasma samples were IgG positive (Guo *et al*., 2020b). Following exposure to the SARS-CoV-2 infection, a humoral immune response is mounted which starts with IgM formation after 3-7 days, indicating an acute or continuing infection and those levels peak after three weeks (Xiao *et al*., 2020). Increased IgG levels may be detected 14 days after symptom onset (Guo *et al*., 2020a). In fact, the antibody dynamics in COVID-19 are quite similar to those in other viral infections, with IgG levels increasing when IgM levels start to decrease (Zhou *et al*., 2020c). It is well known that IL-6 may drive IgG formation by B-cells (Maeda *et al*., 2010). Increased IgG titers are more prevalent in severe than in mild clinical status, especially in female patients (Zeng *et al*.). As such, the association between positive IgG titers and ACE2 and sRAGE could be explained by the increased severity of illness.

## Data Availability

The dataset generated during and/or analyzed during the current study will be available from MM upon reasonable request and once the dataset has been fully exploited by the authors.

## Acknowledgment

We thank the staff of Al-Sadr Teaching Hospital and Al-Amal Specialized Hospital for Communicable Diseases in Najaf governorate-Iraq for their help in the collection of samples. We also thank the high-skilled staff of the hospital’s internal labs, especially Mr. Murtadha Al-Hilo, for their help in the estimation of biomarkers levels.

## Author’s contributions

HAJ recruited the patients and collected blood samples. HAJ and HAH measured the serum biomarkers. MM performed the statistical analysis. All authors collaborated in the analysis design, the discussion of the findings, the drafting and editing of the manuscript, and the final version of the manuscript.

## References

1. Abiri, B. & Vafa, M., 2020. Effects of vitamin D and/or magnesium supplementation on mood, serum levels of BDNF, inflammatory biomarkers, and SIRT1 in obese women: a study protocol for a double-blind, randomized, placebo-controlled trial. Trials, 21, 225.

2. Adams, H.J., Kwee, T.C., Yakar, D., Hope, M.D. & Kwee, R.M., 2020. Chest CT imaging signature of coronavirus disease 2019 infection: in pursuit of the scientific evidence. Chest, 158, 1885–1895.

3. Amini Pouya, M., Afshani, S.M., Maghsoudi, A.S., Hassani, S. & Mirnia, K., 2020. Classification of the present pharmaceutical agents based on the possible effective mechanism on the COVID-19 infection. Daru, 1–20.

4. Azkur, A.K., Akdis, M., Azkur, D., Sokolowska, M., Van De Veen, W., Brüggen, M.C., O’mahony, L., Gao, Y., Nadeau, K. & Akdis, C.A., 2020. Immune response to SARS-CoV-2 and mechanisms of immunopathological changes in COVID-19. Allergy, 75, 1564–1581.

5. Benjamini, Y. & Hochberg, Y., 1995. Controlling the False Discovery Rate: A Practical and Powerful Approach to Multiple Testing. Journal of the Royal Statistical Society: Series B (Methodological), 57, 289–300.

6. Berry, C.T., May, M.J. & Freedman, B.D., 2018. STIM- and Orai-mediated calcium entry controls NF-κB activity and function in lymphocytes. Cell calcium, 74, 131–143.

7. Booth, C.M., 2003. Clinical Features and Short-term Outcomes of 144 Patients With SARS in the Greater Toronto Area. JAMA, 289, 2801.

8. Brest, P., Mograbi, B., Hofman, P. & Milano, G., 2021. More light on cancer and COVID-19 reciprocal interaction. British Journal of Cancer.

9. Cappellini, F., Brivio, R., Casati, M., Cavallero, A., Contro, E. & Brambilla, P., 2020. Low levels of total and ionized calcium in blood of COVID-19 patients. Clinical Chemistry and Laboratory Medicine (CCLM), 58, e171–e173.

10. Chen, N., Zhou, M., Dong, X., Qu, J., Gong, F., Han, Y., Qiu, Y., Wang, J., Liu, Y. & Wei, Y., 2020. Epidemiological and clinical characteristics of 99 cases of 2019 novel coronavirus pneumonia in Wuhan, China: a descriptive study. The lancet, 395, 507–513.

11. Chen, S., Zhang, Z., Wu, Y., Shi, Q., Yan, H., Mei, N., Tolleson, W.H. & Guo, L., 2015. Endoplasmic Reticulum Stress and Store-Operated Calcium Entry Contribute to Usnic Acid-Induced Toxicity in Hepatic Cells. Toxicological sciences : an official journal of the Society of Toxicology, 146, 116–126.

12. Chen, X., Cao, R. & Zhong, W., 2019. Host Calcium Channels and Pumps in Viral Infections. Cells, 9, 94.

13. Çiçek, G., Açıkgoz, S.K., Yayla, Ç., Kundi, H. & İleri, M., 2016. Magnesium as a predictor of acute stent thrombosis in patients with ST-segment elevation myocardial infarction who underwent primary angioplasty. Coron Artery Dis, 27, 47–51.

14. Civitelli, R. & Ziambaras, K., 2011. Calcium and phosphate homeostasis: concerted interplay of new regulators. J Endocrinol Invest, 34, 3–7.

15. Dai, W.-C., Zhang, H.-W., Yu, J., Xu, H.-J., Chen, H., Luo, S.-P., Zhang, H., Liang, L.-H., Wu, X.-L. & Lei, Y., 2020. CT imaging and differential diagnosis of COVID-19. Canadian Association of Radiologists Journal, 71, 195–200.

16. Deng, B., Zhang, S., Geng, Y., Zhang, Y., Wang, Y., Yao, W., Wen, Y., Cui, W., Zhou, Y. & Gu, Q., 2012. Cytokine and chemokine levels in patients with severe fever with thrombocytopenia syndrome virus. PloS one, 7, e41365.

17. Di Filippo, L., Formenti, A.M., Doga, M., Frara, S., Rovere-Querini, P., Bosi, E., Carlucci, M. & Giustina, A., 2021. Hypocalcemia is a distinctive biochemical feature of hospitalized COVID-19 patients. Endocrine, 71, 9–13.

18. Di Filippo, L., Formenti, A.M., Rovere-Querini, P., Carlucci, M., Conte, C., Ciceri, F., Zangrillo, A. & Giustina, A., 2020. Hypocalcemia is highly prevalent and predicts hospitalization in patients with COVID-19. Endocrine, 68, 475–478.

19. Dozio, E., Sitzia, C., Pistelli, L., Cardani, R., Rigolini, R., Ranucci, M. & Corsi Romanelli, M.M., 2020a. Soluble Receptor for Advanced Glycation End Products and Its Forms in COVID-19 Patients with and without Diabetes Mellitus: A Pilot Study on Their Role as Disease Biomarkers. J Clin Med, 9.

20. Dozio, E., Sitzia, C., Pistelli, L., Cardani, R., Rigolini, R., Ranucci, M. & Corsi Romanelli, M.M., 2020b. Soluble Receptor for Advanced Glycation End Products and Its Forms in COVID-19 Patients with and without Diabetes Mellitus: A Pilot Study on Their Role as Disease Biomarkers. Journal of Clinical Medicine, 9, 3785.

21. Dresselhaus, E.C. & Meffert, M.K., 2019. Cellular Specificity of NF-κB Function in the Nervous System. Frontiers in Immunology, 10.

22. Ekong, U., Zeng, S., Dun, H., Feirt, N., Guo, J., Ippagunta, N., Guarrera, J.V., Lu, Y., Weinberg, A., Qu, W., Ramasamy, R., Schmidt, A.M. & Emond, J.C., 2006. Blockade of the receptor for advanced glycation end products attenuates acetaminophen-induced hepatotoxicity in mice. J Gastroenterol Hepatol, 21, 682–8.

23. Eltzschig, H.K., Bratton, D.L. & Colgan, S.P., 2014. Targeting hypoxia signalling for the treatment of ischaemic and inflammatory diseases. Nature Reviews Drug Discovery, 13, 852–869.

24. Fang, Y., 2020. Fang Y, Zhang H, Xie J, et al. Sensitivity of chest CT for COVID-19: comparison to RT-PCR. Radiology, 200432.

25. Franquet, T., 2011. Imaging of pulmonary viral pneumonia. Radiology. 260(1), 18–39. doi: 10.1148/radiol.11092149. PMID: 21697307.

26. Fung, T.S. & Liu, D.X., 2014. Coronavirus infection, ER stress, apoptosis and innate immunity. Frontiers in Microbiology, 5.

27. Gao, Y., Li, T., Han, M., Li, X., Wu, D., Xu, Y., Zhu, Y., Liu, Y., Wang, X. & Wang, L., 2020. Diagnostic utility of clinical laboratory data determinations for patients with the severe COVID-19. J Med Virol, 92, 791–796.

28. Glowacka, I., Bertram, S., Herzog, P., Pfefferle, S., Steffen, I., Muench, M.O., Simmons, G., Hofmann, H., Kuri, T. & Weber, F., 2010. Differential downregulation of ACE2 by the spike proteins of severe acute respiratory syndrome coronavirus and human coronavirus NL63. Journal of virology, 84, 1198–1205.

29. Görlach, A., Bertram, K., Hudecova, S. & Krizanova, O., 2015. Calcium and ROS: a mutual interplay. Redox biology, 6, 260–271.

30. Gromova, O.A., Torshin, I.Y., Kobalava, Z.D., Sorokina, M.A., Villevalde, S.V., Galochkin, S.A., Gogoleva, I.V., Gracheva, O.N., Grishina, T.R., Gromov, A.N., Egorova, E.Y., Kalacheva, A.G., Malyavskaya, S.I., Meraï, I.A. & Semenov, V.A., 2018. Deficit of Magnesium and States of Hypercoagulation: Intellectual Analysis of Data Obtained From a Sample of Patients Aged 18-50 years From Medical and Preventive Facilities in Russia. Kardiologiia, 58, 22–35.

31. Guan, W.-J., Ni, Z.-Y., Hu, Y., Liang, W.-H., Ou, C.-Q., He, J.-X., Liu, L., Shan, H., Lei, C.-L. & Hui, D.S., 2020a. Clinical characteristics of coronavirus disease 2019 in China. New England journal of medicine, 382, 1708–1720.

32. Guan, W.J., Ni, Z.Y., Hu, Y., Liang, W.H., Ou, C.Q., He, J.X., Liu, L., Shan, H., Lei, C.L., Hui, D.S.C., Du, B., Li, L.J., Zeng, G., Yuen, K.Y., Chen, R.C., Tang, C.L., Wang, T., Chen, P.Y., Xiang, J., Li, S.Y., Wang, J.L., Liang, Z.J., Peng, Y.X., Wei, L., Liu, Y., Hu, Y.H., Peng, P., Wang, J.M., Liu, J.Y., Chen, Z., Li, G., Zheng, Z.J., Qiu, S.Q., Luo, J., Ye, C.J., Zhu, S.Y. & Zhong, N.S., 2020b. Clinical Characteristics of Coronavirus Disease 2019 in China. N Engl J Med, 382, 1708–1720.

33. Guo, L., Ren, L., Yang, S., Xiao, M., Chang, D., Yang, F., Dela Cruz, C.S., Wang, Y., Wu, C. & Xiao, Y., 2020a. Profiling early humoral response to diagnose novel coronavirus disease (COVID-19). Clinical Infectious Diseases, 71, 778–785.

34. Guo, L., Ren, L., Yang, S., Xiao, M., Chang, D., Yang, F., Dela Cruz, C.S., Wang, Y., Wu, C., Xiao, Y., Zhang, L., Han, L., Dang, S., Xu, Y., Yang, Q.W., Xu, S.Y., Zhu, H.D., Xu, Y.C., Jin, Q., Sharma, L., Wang, L. & Wang, J., 2020b. Profiling Early Humoral Response to Diagnose Novel Coronavirus Disease (COVID-19). Clin Infect Dis, 71, 778–785.

35. Güzel, A., Doğan, E., Türkçü, G., Kuyumcu, M., Kaplan, İ., Çelik, F. & Yildirim, Z.B., 2019. Dexmedetomidine and Magnesium Sulfate: A Good Combination Treatment for Acute Lung Injury? J Invest Surg, 32, 331–342.

36. Han, H., Ma, Q., Li, C., Liu, R., Zhao, L., Wang, W., Zhang, P., Liu, X., Gao, G., Liu, F., Jiang, Y., Cheng, X., Zhu, C. & Xia, Y., 2020. Profiling serum cytokines in COVID-19 patients reveals IL-6 and IL-10 are disease severity predictors. Emerging Microbes & Infections, 9, 1123–1130.

37. Hansell, D.M., Bankier, A.A., MacMahon, H., McLoud, T.C., Müller, N.L., Remy, J., 2008. Fleischner Society: glossary of terms for thoracic imaging. Radiology. 246(3), 697–722. doi: 10.1148/radiol.2462070712. Epub 2008 Jan 14. PMID: 18195376.

38. Hoffmann, M., Kleine-Weber, H., Schroeder, S., Krüger, N., Herrler, T., Erichsen, S., Schiergens, T.S., Herrler, G., Wu, N.H., Nitsche, A., Müller, M.A., Drosten, C. & Pöhlmann, S., 2020. SARS-CoV-2 Cell Entry Depends on ACE2 and TMPRSS2 and Is Blocked by a Clinically Proven Protease Inhibitor. Cell, 181, 271-280.e8.

39. Huang, C., Wang, Y., Li, X., Ren, L., Zhao, J., Hu, Y., Zhang, L., Fan, G., Xu, J. & Gu, X., 2020a. Clinical features of patients infected with 2019 novel coronavirus in Wuhan, China. The lancet, 395, 497–506.

40. Huang, C., Wang, Y., Li, X., Ren, L., Zhao, J., Hu, Y., Zhang, L., Fan, G., Xu, J., Gu, X., Cheng, Z., Yu, T., Xia, J., Wei, Y., Wu, W., Xie, X., Yin, W., Li, H., Liu, M., Xiao, Y., Gao, H., Guo, L., Xie, J., Wang, G., Jiang, R., Gao, Z., Jin, Q., Wang, J. & Cao, B., 2020b. Clinical features of patients infected with 2019 novel coronavirus in Wuhan, China. Lancet, 395, 497–506.

41. Huang, J., Cheng, A., Kumar, R., Fang, Y., Chen, G., Zhu, Y. & Lin, S., 2020c. Hypoalbuminemia predicts the outcome of COVID-19 independent of age and co-morbidity. J Med Virol, 92, 2152–2158.

42. Hui, D.S. & Zumla, A., 2019. Severe acute respiratory syndrome: historical, epidemiologic, and clinical features. Infectious Disease Clinics, 33, 869–889.

43. Hyser, J.M. & Estes, M.K., 2015. Pathophysiological Consequences of Calcium-Conducting Viroporins. Annual review of virology, 2, 473–496.

44. Izushi, Y., Teshigawara, K., Liu, K., Wang, D., Wake, H., Takata, K., Yoshino, T., Takahashi, H.K., Mori, S. & Nishibori, M., 2016. Soluble form of the receptor for advanced glycation end-products attenuates inflammatory pathogenesis in a rat model of lipopolysaccharide-induced lung injury. J Pharmacol Sci, 130, 226–34.

45. Jia, H., 2016. Pulmonary angiotensin-converting enzyme 2 (ACE2) and inflammatory lung disease. Shock, 46, 239–248.

46. Julio, C.T.-V., Elvis, C. & Mónica, A.T.-R., 2014. Receptor for AGEs (RAGE) as Mediator of NF-kB Pathway Activation in Neuroinflammation and Oxidative Stress. CNS & Neurological Disorders - Drug Targets, 13, 1615–1626.

47. Kelly, A. & Levine, M.A., 2013. Hypocalcemia in the critically ill patient. J Intensive Care Med, 28, 166–77.

48. Kuba, K., Imai, Y., Ohto-Nakanishi, T. & Penninger, J.M., 2010. Trilogy of ACE2: A peptidase in the renin– angiotensin system, a SARS receptor, and a partner for amino acid transporters. Pharmacology & therapeutics, 128, 119–128.

49. Kwee, T.C. & Kwee, R.M., 2020. Chest CT in COVID-19: What the Radiologist Needs to Know. RadioGraphics, 40, 1848–1865.

50. Lambert, D.W., Yarski, M., Warner, F.J., Thornhill, P., Parkin, E.T., Smith, A.I., Hooper, N.M. & Turner, A.J., 2005. Tumor necrosis factor-alpha convertase (ADAM17) mediates regulated ectodomain shedding of the severe-acute respiratory syndrome-coronavirus (SARS-CoV) receptor, angiotensin-converting enzyme-2 (ACE2). J Biol Chem, 280, 30113–9.

51. Li, G., Fan, Y., Lai, Y., Han, T., Li, Z., Zhou, P., Pan, P., Wang, W., Hu, D. & Liu, X., 2020a. Coronavirus infections and immune responses. Journal of medical virology, 92, 424–432.

52. Li, L., Li, S., Xu, M., Yu, P., Zheng, S., Duan, Z., Liu, J., Chen, Y. & Li, J., 2020b. Risk factors related to hepatic injury in patients with corona virus disease 2019. MedRxiv.

53. Lippi, G., South, A.M. & Henry, B.M., 2020. Electrolyte imbalances in patients with severe coronavirus disease 2019 (COVID-19). Annals of clinical biochemistry, 57, 262–265.

54. Luks, A.M. & Swenson, E.R., 2020. COVID-19 Lung Injury and High-Altitude Pulmonary Edema. A False Equation with Dangerous Implications. Annals of the American Thoracic Society, 17, 918–921.

55. Macaione, V., Aguennouz, M., Rodolico, C., Mazzeo, A., Patti, A., Cannistraci, E., Colantone, L., Di Giorgio, R.M., De Luca, G. & Vita, G., 2007. RAGE-NF-kappaB pathway activation in response to oxidative stress in facioscapulohumeral muscular dystrophy. Acta Neurol Scand, 115, 115–21.

56. Maclaren, G., Fisher, D. & Brodie, D., 2020. Preparing for the Most Critically Ill Patients With COVID-19: The Potential Role of Extracorporeal Membrane Oxygenation. Jama, 323, 1245–1246.

57. Maeda, K., Mehta, H., Drevets, D.A. & Coggeshall, K.M., 2010. IL-6 increases B-cell IgG production in a feed-forward proinflammatory mechanism to skew hematopoiesis and elevate myeloid production. Blood, The Journal of the American Society of Hematology, 115, 4699–4706.

58. Maes, M., 1993 A review on the acute phase response in major depression. Rev Neurosci. 4(4), 407–416. doi: 10.1515/revneuro.1993.4.4.407. PMID: 7506108.

59. Maes, M., Galecki, P., Chang, Y.S. & Berk, M., 2011. A review on the oxidative and nitrosative stress (O&NS) pathways in major depression and their possible contribution to the (neuro)degenerative processes in that illness. Prog Neuropsychopharmacol Biol Psychiatry, 35, 676–92.

60. Mehta, P., Mcauley, D.F., Brown, M., Sanchez, E., Tattersall, R.S. & Manson, J.J., 2020. COVID-19: consider cytokine storm syndromes and immunosuppression. Lancet, 395, 1033–1034.

61. Millet, J.K. & Whittaker, G.R., 2018. Physiological and molecular triggers for SARS-CoV membrane fusion and entry into host cells. Virology, 517, 3–8.

62. Morris, G., Athan, E., Walder, K., Bortolasci, C.C., O’neil, A., Marx, W., Berk, M., Carvalho, A.F., Maes, M. & Puri, B.K., 2020a. Can endolysosomal deacidification and inhibition of autophagy prevent severe COVID-19? Life Sciences, 262, 118541.

63. Morris, G., Bortolasci, C.C., Puri, B.K., Olive, L., Marx, W., O’neil, A., Athan, E., Carvalho, A.F., Maes, M., Walder, K. & Berk, M., 2020b. The pathophysiology of SARS-CoV-2: A suggested model and therapeutic approach. Life sciences, 258, 118166–118166.

64. Moustafa, S.R., Al-Rawi, K.F., Al-Dujaili, A.H., Supasitthumrong, T., Al-Hakeim, H.K. & Maes, M., 2020. The Endogenous Opioid System in Schizophrenia and Treatment Resistant Schizophrenia: Increased Plasma Endomorphin 2, and κ and μ Opioid Receptors are Associated with Interleukin-6.

65. Nagy, B., Jr., Fejes, Z., Szentkereszty, Z., Sütő, R., Várkonyi, I., Ajzner, É., Kappelmayer, J., Papp, Z., Tóth, A. & Fagyas, M., 2021. A dramatic rise in serum ACE2 activity in a critically ill COVID-19 patient. Int J Infect Dis, 103, 412–414.

66. Narula, S., Yusuf, S., Chong, M., Ramasundarahettige, C., Rangarajan, S., Bangdiwala, S.I., Van Eikels, M., Leineweber, K., Wu, A. & Pigeyre, M., 2020. Plasma ACE2 and risk of death or cardiometabolic diseases: a case-cohort analysis. The Lancet, 396, 968–976.

67. Nielsen, F.H., 2018. Magnesium deficiency and increased inflammation: current perspectives. Journal of inflammation research, 11, 25.

68. Nieto-Torres, J.L., Verdiá-Báguena, C., Jimenez-Guardeño, J.M., Regla-Nava, J.A., Castaño-Rodriguez, C., Fernandez-Delgado, R., Torres, J., Aguilella, V.M. & Enjuanes, L., 2015. Severe acute respiratory syndrome coronavirus E protein transports calcium ions and activates the NLRP3 inflammasome. Virology, 485, 330–339.

69. Nunes, P. & Demaurex, N., 2014. Redox regulation of store-operated Ca2+ entry. Antioxidants & redox signaling, 21, 915–932.

70. Oczypok, E.A., Perkins, T.N. & Oury, T.D., 2017. All the “RAGE” in lung disease: The receptor for advanced glycation endproducts (RAGE) is a major mediator of pulmonary inflammatory responses. Paediatr Respir Rev, 23, 40–49.

71. Oliveira, B.A., Oliveira, L.C., Sabino, E.C. & Okay, T.S., 2020. SARS-CoV-2 and the COVID-19 disease: a mini review on diagnostic methods. Rev Inst Med Trop Sao Paulo, 62, e44.

72. Oudit, G.Y. & Pfeffer, M.A., 2020. Plasma angiotensin-converting enzyme 2: novel biomarker in heart failure with implications for COVID-19. European heart journal, 41, 1818–1820.

73. Paar, M., Rossmann, C., Nusshold, C., Wagner, T., Schlagenhauf, A., Leschnik, B., Oettl, K., Koestenberger, M., Cvirn, G. & Hallström, S., 2017. Anticoagulant action of low, physiologic, and high albumin levels in whole blood. PLoS One, 12, e0182997.

74. Pal, R., Ram, S., Zohmangaihi, D., Biswas, I., Suri, V., Yaddanapudi, L.N., Malhotra, P., Soni, S.L., Puri, G.D., Bhalla, A. & Bhadada, S.K., 2020. High Prevalence of Hypocalcemia in Non-severe COVID-19 Patients: A Retrospective Case-Control Study. Front Med (Lausanne), 7, 590805.

75. Pan, F., Ye, T., Sun, P., Gui, S., Liang, B., Li, L., Zheng, D., Wang, J., Hesketh, R.L. & Yang, L., 2020. Time course of lung changes on chest CT during recovery from 2019 novel coronavirus (COVID-19) pneumonia. Radiology.

76. Patel, S.K., Juno, J.A., Lee, W.S., Wragg, K.M., Hogarth, P.M., Kent, S.J. & Burrell, L.M., 2021. Plasma ACE2 activity is persistently elevated following SARS-CoV-2 infection: implications for COVID-19 pathogenesis and consequences. European Respiratory Journal.

77. Qin, C., Zhou, L., Hu, Z., Zhang, S., Yang, S., Tao, Y., Xie, C., Ma, K., Shang, K., Wang, W. & Tian, D.S., 2020. Dysregulation of Immune Response in Patients With Coronavirus 2019 (COVID-19) in Wuhan, China. Clin Infect Dis, 71, 762–768.

78. Quilliot, D., Bonsack, O., Jaussaud, R. & Mazur, A., 2020. Dysmagnesemia in Covid-19 cohort patients: prevalence and associated factors. Magnesium Research, 33, 114–122.

79. Romani, A. & Scarpa, A., 1992. Regulation of cell magnesium. Archives of biochemistry and biophysics, 298, 1–12.

80. Ronit, A., Kirkegaard-Klitbo, D.M., Dohlmann, T.L., Lundgren, J., Sabin, C.A., Phillips, A.N., Nordestgaard, B.G. & Afzal, S., 2020. Plasma albumin and incident cardiovascular disease: results from the CGPS and an updated meta-analysis. Arteriosclerosis, thrombosis, and vascular biology, 40, 473–482.

81. Ruan, Q., Yang, K., Wang, W., Jiang, L. & Song, J., 2020. Clinical predictors of mortality due to COVID-19 based on an analysis of data of 150 patients from Wuhan, China. Intensive Care Med, 46, 846–848.

82. Sadhukhan, P., Ugurlu, M.T. & Hoque, M.O., 2020. Effect of COVID-19 on Lungs: Focusing on Prospective Malignant Phenotypes. Cancers, 12, 3822.

83. Shenoy, N., Luchtel, R. & Gulani, P., 2020. Considerations for target oxygen saturation in COVID-19 patients: are we under-shooting? BMC Med, 18, 260.

84. Sheu, J.R., Hsiao, G., Shen, M.Y., Lee, Y.M. & Yen, M.H., 2003. Antithrombotic effects of magnesium sulfate in in vivo experiments. Int J Hematol, 77, 414–9.

85. Soeters, P.B., Wolfe, R.R. & Shenkin, A., 2019. Hypoalbuminemia: pathogenesis and clinical significance. Journal of Parenteral and Enteral Nutrition, 43, 181–193.

86. Song, J.-W., Zhang, C., Fan, X., Meng, F.-P., Xu, Z., Xia, P., Cao, W.-J., Yang, T., Dai, X.-P. & Wang, S.-Y., 2020. Immunological and inflammatory profiles in mild and severe cases of COVID-19. Nature communications, 11, 1–10.

87. Sterenczak, K.A., Willenbrock, S., Barann, M., Klemke, M., Soller, J.T., Eberle, N., Nolte, I., Bullerdiek, J. & Murua Escobar, H., 2009. Cloning, characterisation, and comparative quantitative expression analyses of receptor for advanced glycation end products (RAGE) transcript forms. Gene, 434, 35–42.

88. Straus, M.R., Tang, T., Lai, A.L., Flegel, A., Bidon, M., Freed, J.H., Daniel, S. & Whittaker, G.R., 2019. Ca2+ ions promote fusion of Middle East Respiratory Syndrome coronavirus with host cells and increase infectivity. Cold Spring Harbor Laboratory.

89. Sukkar, M.B., Wood, L.G., Tooze, M., Simpson, J.L., Mcdonald, V.M., Gibson, P.G. & Wark, P.A., 2012. Soluble RAGE is deficient in neutrophilic asthma and COPD. Eur Respir J, 39, 721–9.

90. Sun, J.K., Zhang, W.H., Zou, L., Liu, Y., Li, J.J., Kan, X.H., Dai, L., Shi, Q.K., Yuan, S.T., Yu, W.K., Xu, H.Y., Gu, W. & Qi, J.W., 2020. Serum calcium as a biomarker of clinical severity and prognosis in patients with coronavirus disease 2019. Aging (Albany NY), 12, 11287–11295.

91. Tetro, J.A., 2020. Is COVID-19 receiving ADE from other coronaviruses? Microbes and infection, 22, 72–73.

92. Thachil, J., Tang, N., Gando, S., Falanga, A., Cattaneo, M., Levi, M., Clark, C. & Iba, T., 2020. ISTH interim guidance on recognition and management of coagulopathy in COVID-19. Journal of Thrombosis and Haemostasis, 18, 1023–1026.

93. Uchida, T., Shirasawa, M., Ware, L.B., Kojima, K., Hata, Y., Makita, K., Mednick, G., Matthay, Z.A. & Matthay, M.A., 2006. Receptor for advanced glycation end-products is a marker of type I cell injury in acute lung injury. American journal of respiratory and critical care medicine, 173, 1008–1015.

94. Van Zoelen, M.a.D., Achouiti, A. & Van Der Poll, T., 2011. The role of receptor for advanced glycation endproducts (RAGE) in infection. Critical Care, 15, 208.

95. Vistoli, G., De Maddis, D., Cipak, A., Zarkovic, N., Carini, M. & Aldini, G., 2013. Advanced glycoxidation and lipoxidation end products (AGEs and ALEs): an overview of their mechanisms of formation. Free Radic Res, 47 Suppl 1, 3–27.

96. Vlachakis, D., Papakonstantinou, E., Mitsis, T., Pierouli, K., Diakou, I., Chrousos, G., Bacopoulou, F.J.F. & Toxicology, C., 2020. Molecular mechanisms of the novel coronavirus SARS-CoV-2 and potential anti-COVID19 pharmacological targets since the outbreak of the pandemic. 111805.

97. Walls, A.C., Park, Y.-J., Tortorici, M.A., Wall, A., Mcguire, A.T. & Veesler, D., 2020. Structure, function, and antigenicity of the SARS-CoV-2 spike glycoprotein. Cell, 181, 281-292. e6.

98. Wang, D., Hu, B., Hu, C., Zhu, F., Liu, X., Zhang, J., Wang, B., Xiang, H., Cheng, Z. & Xiong, Y., 2020. Clinical characteristics of 138 hospitalized patients with 2019 novel coronavirus–infected pneumonia in Wuhan, China. Jama, 323, 1061–1069.

99. Wang, Y. & Liu, L., 2016. The Membrane Protein of Severe Acute Respiratory Syndrome Coronavirus Functions as a Novel Cytosolic Pathogen-Associated Molecular Pattern To Promote Beta Interferon Induction via a Toll-Like-Receptor-Related TRAF3-Independent Mechanism. mBio, 7, e01872–15.

100. Wu, C., Chen, X., Cai, Y., Xia, J., Zhou, X., Xu, S., Huang, H., Zhang, L., Zhou, X., Du, C., Zhang, Y., Song, J., Wang, S., Chao, Y., Yang, Z., Xu, J., Zhou, X., Chen, D., Xiong, W., Xu, L., Zhou, F., Jiang, J., Bai, C., Zheng, J. & Song, Y., 2020. Risk Factors Associated With Acute Respiratory Distress Syndrome and Death in Patients With Coronavirus Disease 2019 Pneumonia in Wuhan, China. JAMA Intern Med, 180, 934–943.

101. Xiao, A.T., Gao, C. & Zhang, S., 2020. Profile of specific antibodies to SARS-CoV-2: The first report. J Infect, 81, 147–178.

102. Xu, Z., Shi, L., Wang, Y., Zhang, J., Huang, L., Zhang, C., Liu, S., Zhao, P., Liu, H. & Zhu, L., 2020. Pathological findings of COVID-19 associated with acute respiratory distress syndrome. The Lancet respiratory medicine, 8, 420–422.

103. Yalcin Kehribar, D., Cihangiroglu, M., Sehmen, E., Avci, B., Capraz, A., Yildirim Bilgin, A., Gunaydin, C. & Ozgen, M., 2021. The receptor for advanced glycation end product (RAGE) pathway in COVID-19. Biomarkers, 26, 114–118.

104. Yang, C., Ma, X., Wu, J., Han, J., Zheng, Z., Duan, H., Liu, Q., Wu, C., Dong, Y. & Dong, L., 2021. Low serum calcium and phosphorus and their clinical performance in detecting COVID-19 patients. J Med Virol, 93, 1639–1651.

105. Yang, W.I., Lee, D., Lee, D.L., Hong, S.Y., Lee, S.H., Kang, S.M., Choi, D.H., Jang, Y., Kim, S.H. & Park, S., 2014. Blocking the receptor for advanced glycation end product activation attenuates autoimmune myocarditis. Circ J, 78, 1197–205.

106. Yang, X., Yu, Y., Xu, J., Shu, H., Liu, H., Wu, Y., Zhang, L., Yu, Z., Fang, M. & Yu, T., 2020. Clinical course and outcomes of critically ill patients with SARS-CoV-2 pneumonia in Wuhan, China: a single-centered, retrospective, observational study. The Lancet Respiratory Medicine, 8, 475–481.

107. Ye, Q., Wang, B. & Mao, J., 2020. The pathogenesis and treatment of the ‘Cytokine Storm’ in COVID-19. J Infect, 80, 607–613.

108. Zainol Rashid, Z., Othman, S.N., Abdul Samat, M.N., Ali, U.K. & Wong, K.K., 2020. Diagnostic performance of COVID-19 serology assays. Malays J Pathol, 42, 13–21.

109. Zeng, F., Dai, C., Cai, P., Wang, J. & Xu, L., Jianyu Li1, Guoyun Hu1, LW (2020). A comparison study of SARS-CoV-2 IgG antibody between male and female COVID-19 patients: a possible reason underlying different outcome between gender. BMJ, February 2019, 1–13.

110. Zhang, C., Wang, J., Ma, X., Wang, W., Zhao, B., Chen, Y., Chen, C. & Bihl, J.C., 2018. ACE2-EPC-EXs protect ageing ECs against hypoxia/reoxygenation-induced injury through the miR- 18a/Nox2/ROS pathway. J Cell Mol Med, 22, 1873–1882.

111. Zhang, F., Su, X., Huang, G., Xin, X.F., Cao, E.H., Shi, Y. & Song, Y., 2017. sRAGE alleviates neutrophilic asthma by blocking HMGB1/RAGE signalling in airway dendritic cells. Sci Rep, 7, 14268.

112. Zhang, H.-W., Yu, J., Xu, H.-J., Lei, Y., Pu, Z.-H., Dai, W.-C., Lin, F., Wang, Y.-L., Wu, X.-L. & Liu, L.-H., 2020a. Corona virus international public health emergencies: implications for radiology management. Academic radiology, 27, 463–467.

113. Zhang, H., Tasaka, S., Shiraishi, Y., Fukunaga, K., Yamada, W., Seki, H., Ogawa, Y., Miyamoto, K., Nakano, Y., Hasegawa, N., Miyasho, T., Maruyama, I. & Ishizaka, A., 2008a. Role of soluble receptor for advanced glycation end products on endotoxin-induced lung injury. Am J Respir Crit Care Med, 178, 356–62.

114. Zhang, I.X., Ren, J., Vadrevu, S., Raghavan, M. & Satin, L.S., 2020b. ER stress increases store-operated Ca(2+) entry (SOCE) and augments basal insulin secretion in pancreatic beta cells. J Biol Chem, 295, 5685–5700.

115. Zhang, L., Bukulin, M., Kojro, E., Roth, A., Metz, V.V., Fahrenholz, F., Nawroth, P.P., Bierhaus, A. & Postina, R., 2008b. Receptor for advanced glycation end products is subjected to protein ectodomain shedding by metalloproteinases. J Biol Chem, 283, 35507–16.

116. Zhang, Y., Zheng, L., Liu, L., Zhao, M., Xiao, J. & Zhao, Q., 2020c. Liver impairment in COVID-19 patients: A retrospective analysis of 115 cases from a single centre in Wuhan city, China. Liver Int, 40, 2095–2103.

117. Zheltova, A.A., Kharitonova, M.V., Iezhitsa, I.N. & Spasov, A.A., 2016. Magnesium deficiency and oxidative stress: an update. BioMedicine, 6.

118. Zhou, F., Yu, T., Du, R., Fan, G., Liu, Y., Liu, Z., Xiang, J., Wang, Y., Song, B. & Gu, X., 2020a. Clinical course and risk factors for mortality of adult inpatients with COVID-19 in Wuhan, China: a retrospective cohort study. The lancet, 395, 1054–1062.

119. Zhou, F., Yu, T., Du, R., Fan, G., Liu, Y., Liu, Z., Xiang, J., Wang, Y., Song, B., Gu, X., Guan, L., Wei, Y., Li, H., Wu, X., Xu, J., Tu, S., Zhang, Y., Chen, H. & Cao, B., 2020b. Clinical course and risk factors for mortality of adult inpatients with COVID-19 in Wuhan, China: a retrospective cohort study. Lancet, 395, 1054–1062.

120. Zhou, P., Yang, X.-L., Wang, X.-G., Hu, B., Zhang, L., Zhang, W., Si, H.-R., Zhu, Y., Li, B. & Huang, C.-L., 2020c. A pneumonia outbreak associated with a new coronavirus of probable bat origin. nature, 579, 270–273.

121. Zhou, Y., Frey, T.K. & Yang, J.J., 2009. Viral calciomics: interplays between Ca2+ and virus. Cell calcium, 46, 1–17.

122. Zhu, Z., Zhang, Z., Chen, W., Cai, Z., Ge, X., Zhu, H., Jiang, T., Tan, W. & Peng, Y., 2018. Predicting the receptor-binding domain usage of the coronavirus based on kmer frequency on spike protein. Infection, Genetics and Evolution, 61, 183.

